# Analyzing the amplification of Auditory sensation in visually impaired individuals and comparison with normal individuals

**DOI:** 10.1101/2023.03.10.23287122

**Authors:** Dev Himanshubhai Desai

## Abstract

**Background:** It is commonly believed that if a person loses one of their basic senses like sight or hearing, other senses will be amplified to compensate the loss. It has been proven that this amplification does indeed help the person up to an extent, but this increase in the senses as compared to other individuals, who have all senses intact, has not been quantified.

**Aim:**

1. To estimate and compare Auditory Acuity between visually impaired and normal people
2. To calculate the amount of amplification in the Auditory sense in the visually impaired individuals in compared to Normal Individuals

**Methodology:** A Cross Sectional Case-control Pilot Experimental Study was carried out using a method that has been developed by the authors, wherein a pure tone sound at a specific intensity was brought inwards towards the subject from multiple directions and the distance was measured where the participant could hear the sound and was noted. The same experiment was carried out in Normal Individuals with normal 6/6 vision and these measurements were compared. Positon of nose in respect to the direction of sound source was used to denote the results in angles made by the two. A pure tone of 256 Hz, of 30db was used as the sound source and the distance from the subject was measured in meters.

**Results:** Out of total 60 individuals in the study, 45 had some degree of blindness. In both groups, the highest sensitivity is at 0° followed by 45°-315°, then comes 90°-270° and 135°-215° to put nose position of 180° at least amount of sensitivity. A 10% amplification in the distance is present between any and all type of visually impaired individuals and normal individuals whereas totally blind individuals have around 20% amplification compared to normal individuals in the distance to appreciate the sound.

**Conclusion:** A visible difference can be seen in the distance where a total blind person can hear, which is higher than the distance at which a normal individual can hear. High loss of vision, low loss of vision and one eye blind individuals come in the range between these two. Although, the results are not statistically significant, they are clinically present, and a with a larger sample size, a better assessment of this amplification can be done.

## Introduction

It is a proven age-old belief that out of all our senses, i.e. vision, touch, smell, taste and hearing, vision is the one most used by humans. (1) It almost overrides the other senses and has been the greatest tool in our survival and the path of evolution.

Loss of one of the basic senses results in a lot of difficulties for the person, but the toughest loss is that of vision (2) There is a high prevalence of blindness in the country, especially in children. (3) Many programs like Vision 2020 have been implemented by the WHO to reduce the burden of blindness in the world. (4)

It is commonly believed that if a person loses one of their basic senses like sight or hearing, other senses will be amplified to compensate the loss and restore as much functionality as possible. This fact is well documented and known to almost everyone, and yet, there is no proper documentation on how much do the other senses amplify when one sense is lost. (5) (6) (7)Many papers and individuals attribute this to an entity in neurology called “Cortical Plasticity” (8), which means that when one part of the brain responsible for a particular sense becomes less functional due to loss of that particular sense, it will convert itself to a new sense, thereby giving that new sense more neurons as compared to other individuals. This amplifies the sense. But, it is also believed that Cortical Plasticity of any neuron to convert and work for something else is only true for neurons in its vicinity. (9) (10)

This phenomenon of amplification is well documented for blindness i.e. loss of vision, that auditory senses amplify to a great extent so that functionality can be somewhat restored (11), but there are few documentations or studies which actually demonstrate HOW MUCH the amplification is and if the amplification is present, does it correlate to the amount of loss of vision present in the individual, or to the amount of time the individual has been visually impaired or both. (12) (13) (14) (15) (16)

It is important to understand the degree of amplification present to understand how much functionality is regained or to help the individual better. (17)It is also important to understand the amount of amplification, since based on the given hypothesis, Cortical Plasticity, the centers of hearing and vision must be close to each other, whereas in reality, the Center of Vision is in the occipital lobe (18)and the Center of Hearing is in the Temporal Lobe (19), which are not close by, but still amplification is observed.

Here, a new methodology is being used, to quantify sensitivity of auditory sense so that it can be compared with normal individual.

## Methodology

A Cross Sectional Case-control Experimental Study was carried using a method which has been developed by the authors, where a pure tone sound at a specific intensity was brought inwards towards the subject from multiple directions and the distance was measured where the participant could hear the sound and was noted. The same experiment was carried out in Normal Individuals with normal 6/6 vision and these measurements were compared.

### Ethical consideration

The research protocol was approved by relevant institutional review boards or ethics committees and that all human participants gave written informed consent, and the authors have no conflict of interest to declare. The Authors have not received any funding from anyone for this research project and have no financial dilemma.

- Inclusion Criteria: -
  - Participants who were willing to take part in the study and willing to give consent
  - Participants whose Rinne, Weber and Absolute Bone conduction test were Normal
  - For the Case group,
    - Who has been certified as Blind by the Disability agency
  - For the Control group,
    - Participants who have 6/6 vision confirmed by a certified ophthalmologist in the last 4 weeks

- Exclusion Criteria: -
  - Participants not willing to give consent for the experiment
  - Participants whose Rinne, Weber and Absolute Bone Conduction tests were abnormal.

A pure tone of 256Hz and Intensity of 30db was used in a be soundproof room for both visually impaired individuals and Normal vision individuals. The distance was mentioned in different directions and participants’ nose was taken as a reference point to measure the direction of the individual when recording the data (the angle made by a hypothetical line from passing from nose and the direction the sound is coming from). These angles were analyzed to depict various directions. The distance from where the subject could hear the sound was measured in meters from the subjects’ head.

This data was analyzed using Excel and SPSS and complex advance analysis was carried out trying to simplify the gross amount of data. Averages, Standard deviation (St.dev.), Standard error of Mean(SEM) and Confidence interval 95 (CI95) were various statistical tools used for analysis along with charts and graphs.

## Results

## Discussion

Total of 60 individuals were taken for this prototype study in which 15 were control group with 6/6 vision and the rest 45 were Visually impaired individuals with variable degree of blindness depicted in the certificate of disability awarded to them as seen in Table 1.

**Table 1:-.** Distribution of Individuals

| Table 1 :- Distribution of Individuals |  |  |  |  |
| --- | --- | --- | --- | --- |
| Total participants in the study :- 60 |  |  |  |  |
| Normal Vision Individuals | All types Of Blind Individuals |  |  |  |
| 15 | 45 |  |  |  |
|  | High Degree of Blindness | Low Degree of Blindness | One eye Blindness | Total Blindness |
|  | 9 | 14 | 2 | 20 |

Table 2 gives a broad sense of the results. Averages in different groups with various amount of blindness can be appreciated. It can be also be noted that Totally Blind individuals have the highest amount of average distance from where they can appreciate the sound source, showing their high acuity for auditory stimulus. And compared to a normal individual, it seems to be higher in every direction. The results are similar to what has been documented before (15) (14) (17) (16)

**Table 2:-.** Distribution of Participants and their preliminary results

| Table 2:- Distribution of Participants and their preliminary results |  |  |  |  |  |  |  |  |  |  |
| --- | --- | --- | --- | --- | --- | --- | --- | --- | --- | --- |
|  | Sample Size |  | 0° | 45° | 90° | 135° | 180° | 215° | 270° | 315° |
| Normal individuals | 15 | Average | 1.79 | 1.70 | 1.53 | 1.41 | 1.31 | 1.36 | 1.46 | 1.63 |
|  |  | St.dev. | 0.16 | 0.17 | 0.21 | 0.23 | 0.22 | 0.23 | 0.23 | 0.20 |
|  |  | SEM | 0.04 | 0.04 | 0.05 | 0.06 | 0.06 | 0.06 | 0.06 | 0.05 |
|  |  | CI95 | 0.08 | 0.09 | 0.11 | 0.12 | 0.11 | 0.11 | 0.12 | 0.10 |
| High Degree of blindness | 9 | Average | 1.87 | 1.79 | 1.63 | 1.58 | 1.47 | 1.54 | 1.61 | 1.71 |
|  |  | St.dev. | 0.17 | 0.29 | 0.45 | 0.47 | 0.55 | 0.51 | 0.45 | 0.35 |
|  |  | SEM | 0.06 | 0.10 | 0.15 | 0.16 | 0.18 | 0.17 | 0.15 | 0.12 |
|  |  | CI95 | 0.11 | 0.19 | 0.29 | 0.31 | 0.36 | 0.33 | 0.29 | 0.23 |
| Low degree | 14 | Average | 1.83 | 1.69 | 1.64 | 1.49 | 1.36 | 1.41 | 1.55 | 1.66 |
| of blindness |  | St-dev· | 0·30 | 0·36 | 0·41 | 0·51 | 0·54 | 0·53 | 0·50 | 0·39 |
|  |  | SEM | 0·08 | 0·10 | 0·11 | 0·14 | 0·15 | 0·14 | 0·13 | 0·10 |
|  |  | CI95 | 0·16 | 0·19 | 0·21 | 0·27 | 0·29 | 0·28 | 0·26 | 0·20 |
| One eye<br>Blind | 2 | Average | 1·78 | 1·75 | 1·70 | 1·47 | 1·40 | 1·45 | 1·65 | 1·73 |
|  |  | St-dev· | 0·32 | 0·35 | 0·42 | 0·76 | 0·85 | 0·78 | 0·49 | 0·39 |
|  |  | SEM | 0·23 | 0·25 | 0·30 | 0·54 | 0·60 | 0·55 | 0·35 | 0·27 |
|  |  | CI95 | 0·44 | 0·49 | 0·59 | 1·05 | 1·18 | 1·08 | 0·69 | 0·54 |
| Total Blind | 20 | Average | 2·27 | 1·87 | 1·85 | 1·72 | 1·63 | 1·66 | 1·77 | 1·95 |
|  |  | St-dev· | 1·96 | 0·21 | 0·25 | 0·43 | 0·51 | 0·48 | 0·40 | 0·11 |
|  |  | SEM | 0·44 | 0·05 | 0·06 | 0·10 | 0·11 | 0·11 | 0·09 | 0·02 |
|  |  | CI95 | 0·86 | 0·09 | 0·11 | 0·19 | 0·22 | 0·21 | 0·18 | 0·05 |
| Sum of all<br>Blind<br>individuals | 45 | Average | 2·02 | 1·79 | 1·73 | 1·61 | 1·50 | 1·55 | 1·66 | 1·80 |
|  |  | St-dev· | 1·32 | 0·29 | 0·36 | 0·47 | 0·53 | 0·50 | 0·44 | 0·30 |
|  |  | SEM | 0·20 | 0·04 | 0·05 | 0·07 | 0·08 | 0·08 | 0·07 | 0·05 |
|  |  | CI95 | 0·39 | 0·08 | 0·10 | 0·14 | 0·16 | 0·15 | 0·13 | 0·09 |

It becomes pretty evident with the help of Figure 1 that almost in every direction as seen in Table 2, totally blind individuals can appreciate the sound at the highest average distance and is proof enough to state that they have a higher auditory acuity and sensitivity for sound impulses. Following the totally blind individuals, are individuals with high degree of blindness. It can be also seen that one eye blind and low degree blind individuals are almost similar yet higher than normal individuals with 6/6 vision. Table 3 also shows a positive correlation (r=0.28) between the time spent with the disability and the amount of amplification in sharpness which provides further proof that auditory acuity and the sensitivity to identify and appreciate a sound source is amplified and increased in visually impaired individuals in comparison with normal individuals and out of many factors that might be responsible for it, the amount of time spent with the disability is of maximum significance. (17)

**Table 3.** Correlation

|  |  |
| --- | --- |
| Table 3 – Correlation |  |
| Pearson Correlation Coefficient between the time since blindness and the distance when they appreciate the demo sound | R = 0.288 |

**Figure 1:-.**
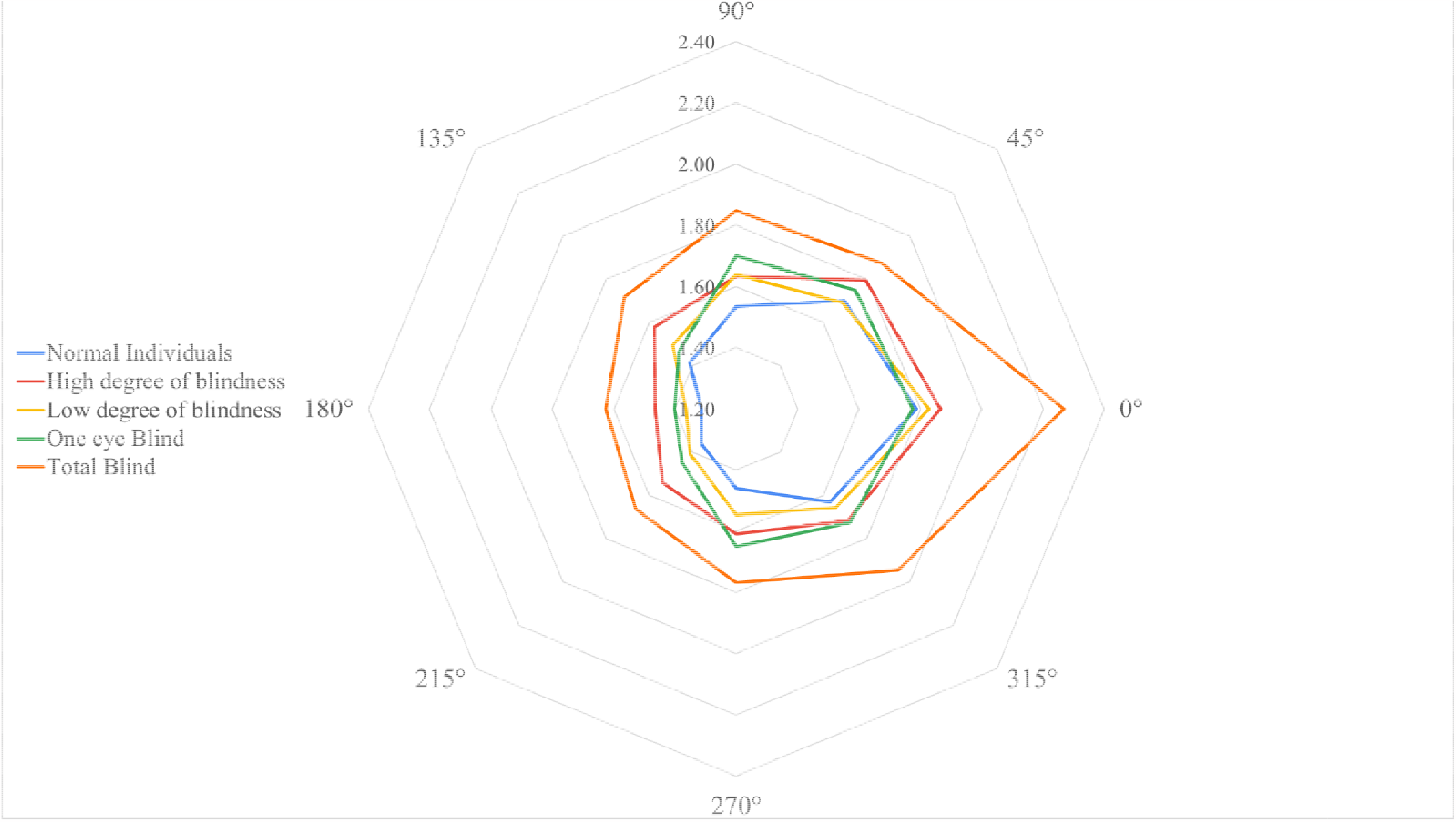
Average Distance to hear the sound for individuals from different groups in different directions

With the help of figure 2, it becomes clearer that the average distance for normal individuals is lower in all directions in compared to all and any type of blindness and a pattern emerges. Figure 1 and figure 2 with the help of table 2 shows pretty clearly that the highest amount of distance to appreciate the sound in any individual is when the nose is at 0°, meaning that surface if helix and antihelix of pinna of both ears is pointing towards the direction of sound source, which is followed by 45° and 315° where the wars are facing the direction of the sound but are not perfectly aligned. Then comes direction of nose at 90° in respect to the sound source meaning that the external auditory canal was directly in line with the sound source followed by 270° when External auditory canal of left ear was in line with the sound source direction. In the end comes nose direction of 135° and 215° where the back part of pinna is facing the direction of sound but the lines are aligned followed by 180° where the imaginary line of direction of incoming sound source will hit the occiput of the subject first.

**Figure 2:-.**
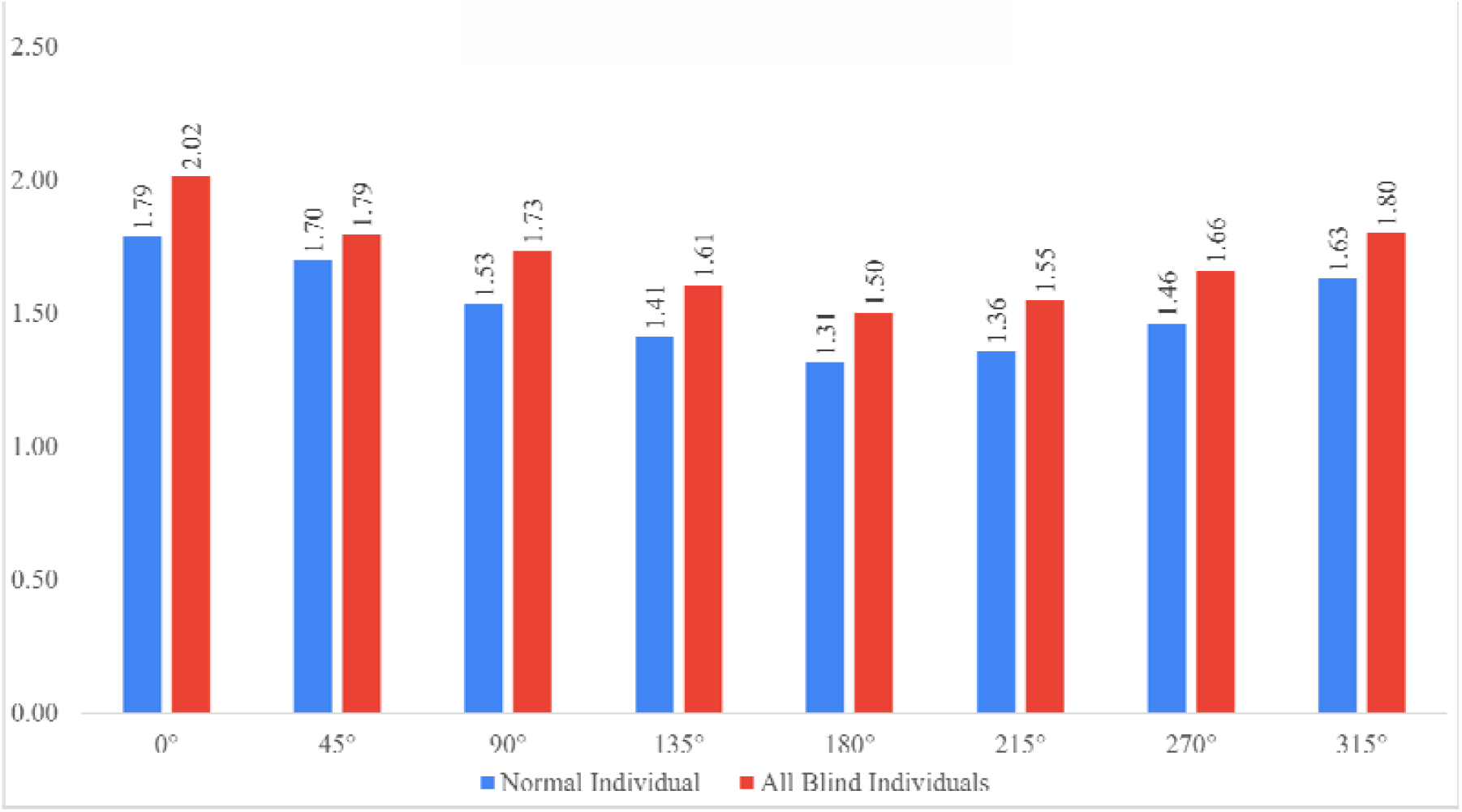
Normal vs Any and all types of blind individuals

Figure 3 is sufficient to demonstrate distance when the sound is appreciated in all and any type of blindness is higher than normal individuals and the sensitivity according to different directions is pretty evident here. A 10% increase can be denoted from Normal individuals in categories of one eye visually impaired and low degree of visual impairment. A 20% increase can be seen in individuals with total visual impairment.

**Figure 3:-.**
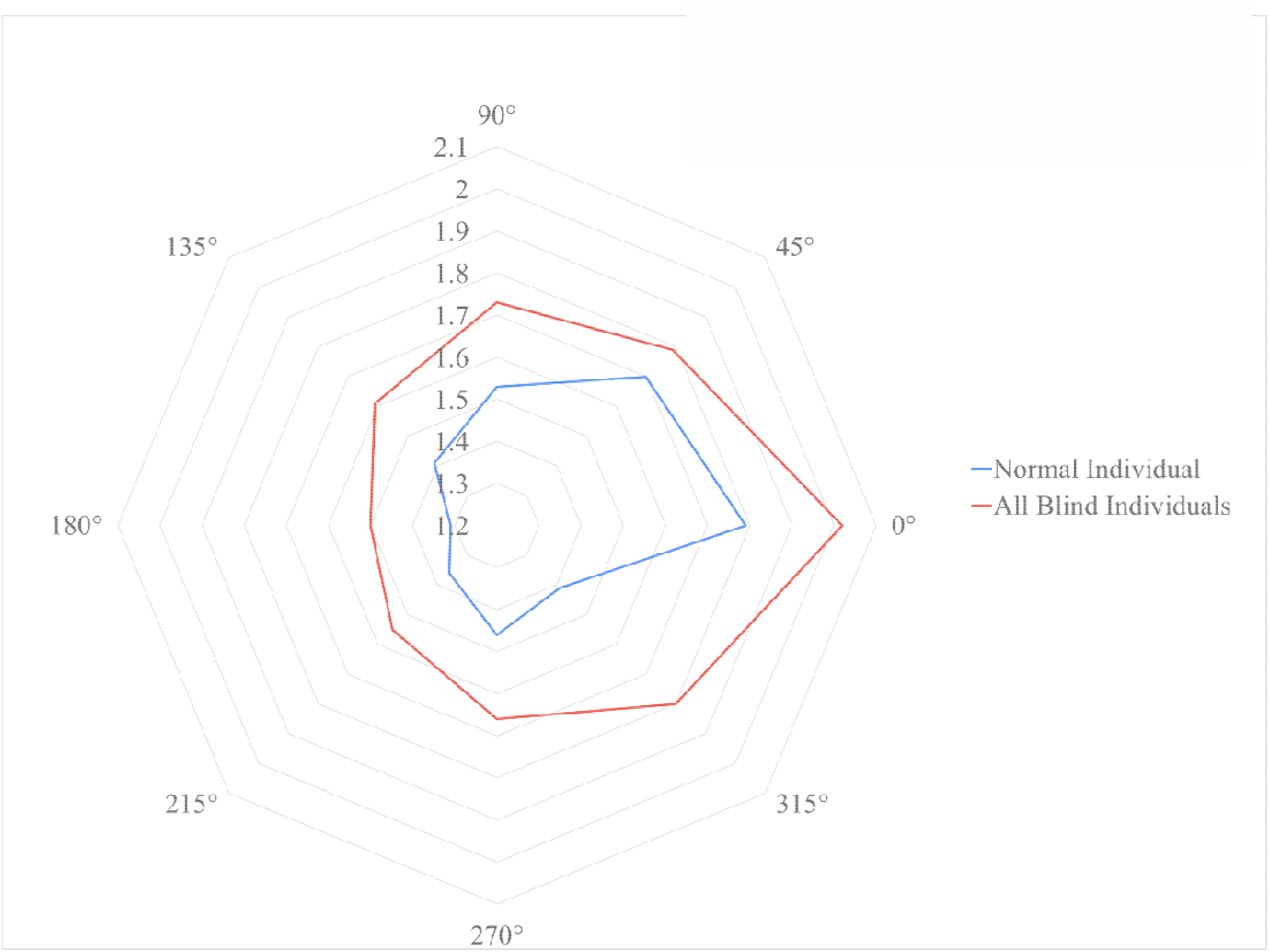
Normal vs Any and all types of blind individuals

It is logical to think that totally blind individuals will have higher amount of sensitivity and hence the distance when they appreciate the demo sound would be higher which should be followed by high degree of blindness. Figure 4 demonstrates that and it also shows that the distance in low degree of blindness and one eye blindness is almost similar and very near to high degree of blindness but it nowhere near to the results of totally blind. It could be that to heighten one sense drastically, complete isolation or loss of one sense might be necessary.

**Figure 4:-.**
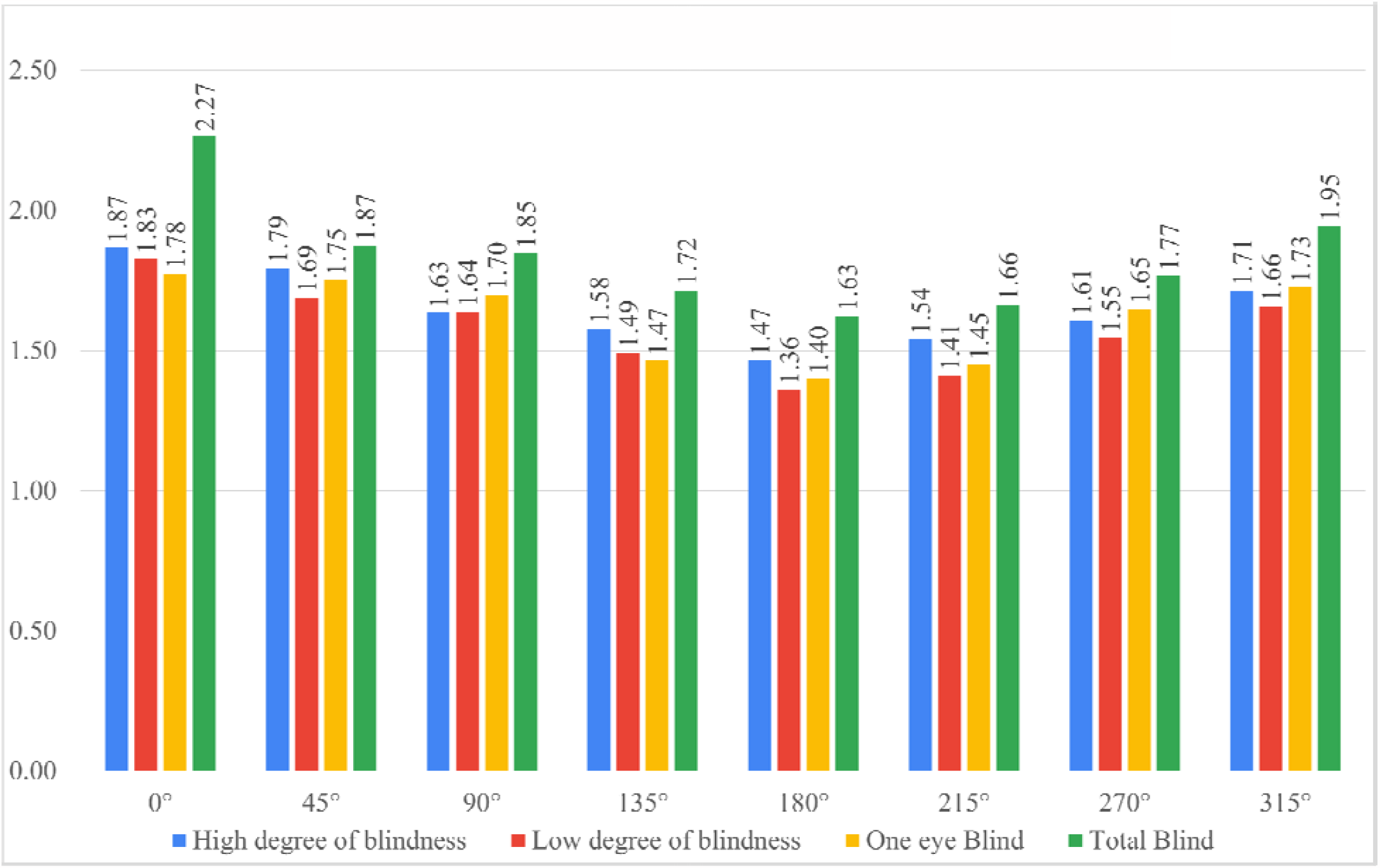
Distance for individuals with different types of blindness

## Conclusion

This results are enough to suffice that in visually impaired individuals a quantifiable difference in amplification in their sensitivity for a sound source and auditory acuity is present which is higher than the sensitivity any normal individual possesses and that sound identification and discrimination is better. Although statistically insignificant, the results are clinically significant and a visible difference is present.

This methodology can be refined and with proper resources, proper funding, and a larger sample size, a more accurate estimation of this amplification can be done.

## Data Availability

All data produced in the present study are available upon reasonable request to the authors

## Acknowledgement

The authors would like to thank Blind School Association, Ahmedabad for providing a place with visually impaired individuals to carry on this research. The authors would also like to thank the following individuals for their efforts along with the authors in data collection. We would like to thank Physiology Department of Smt. NHLMMC and Dr. Anita Verma and Dr. Neeraj Mahajan for their constant support. We would like to thank CA Jaladhi Acharya, Dr. Jugal Shah, Dr. Prahasth Dave, Dr. Deval Patel, Dr. Dwija Raval, Dr. Charmi Shah, Dr. Stuti Chauhan, Dr. Tithi Patel, Dr. Yash Chauhan, Dr. Yash Thumar, Dr. Joey Pindaria, Dr. Dev Pandhi, Dr. Jay Patel, Dr. Dev Andharia, Dr. Nishi Shah, Dr. Kshitish Bhramachari and Dr. Tanvi Sahni for their efforts. We would also like to thank Dr. Dholakia (Sanket Speech and Hearing Clinic) for his help in the development process of the methodology.

